# The Xella Clock: a female-specific epigenetic aging clock optimized for menstrual fluid and endometrial tissue

**DOI:** 10.64898/2026.08.07.26359971

**Authors:** A. Pavuluri, B. Gould, A. Indap, N. Salakh, K. Lacob, A. Dantas, O. Sazonova, J. Ching

## Abstract

The female reproductive system is one of the first major organ systems to show signs of age-related decline, and menopause is associated with increased risk of several diseases, including osteoporosis and cardiovascular disease. Menstrual fluid contains a mixture of blood and endometrial tissue and is a noninvasive biological sample type that has immense potential for diagnostics related to female reproductive aging. However, existing epigenetic aging clocks show limited performance in hormone-dependent tissues such as the endometrium. At Xella Health, we collected menstrual fluid (MF) samples, from a diverse patient cohort (n=66) and quantified genome-wide 5mC methylation levels. We then developed a novel, deep learning-based epigenetic aging clock that is optimized for performance in menstrual fluid and endometrial tissue. Our model, the Xella Clock, outperforms other widely used epigenetic aging clocks at predicting chronological age from MF data and on endometrial tissue. The model is a useful tool for advancing the study of female reproductive aging and can be used to examine associations between endometrial age acceleration and clinical factors.

## INTRODUCTION

Aging is a multifactorial process that results in the gradual decline of biological functions [1]. Over the past few decades, the role of epigenetic alterations in aging has been widely studied, with particular emphasis on DNA methylation (DNAm) [2]. DNAm involves the addition of a methyl group to the 5’ carbon of cytosine, creating 5-methylcytosine at CG dinucleotides (CpG sites) throughout the genome. The addition and removal of methyl groups from CpG sites are integral processes in the regulation of transcription [3], and become increasingly dysregulated as we age due to biology, lifestyle factors, and environmental exposures.

The study of DNAm changes throughout the human lifespan gave rise to the development of the epigenetic aging clock, a powerful tool utilizing machine learning to estimate one’s age from DNAm biomarkers. Horvath’s clock was the first pan-tissue epigenetic clock, proven to accurately and precisely predict age across major organ systems [4]. Moreover, this model identified 353 age-associated CpG sites that are conserved across diverse tissue types; of the CpG sites identified, many were in proximity to genes associated with known hallmarks of aging. Since the publication of the Horvath clock, many other epigenetic clocks have been developed, predicting mortality, lifespan, and healthspan from multiple biological data types [5-7]. While “chronological age” refers to the number of years one has been alive, “biological age” refers to the age predicted by an epigenetic clock, reflecting the physiological and/or molecular state of an individual’s body, capturing their true rate of aging and risk of age-related disease. A person’s biological age may be higher or lower than their chronological age, with a difference indicating the pace of aging of the body in comparison with a reference group. Biological age can also be measured for specific organs or organ systems of the body although there is currently less direct understanding of the implication of such measurements [8]. Actions such as smoking and alcohol consumption may result in accelerated biological aging, while actions such as consistent exercise and healthy diet may result in a biological age lower than the chronological age [9]. Diseases including obesity, cancer, diabetes, and chronic inflammation have also been shown to accelerate biological aging [10, 11].

While epigenetic clocks have proven to be extremely powerful tools for the field of longevity, one major flaw in these tools is that they overlook the highly sex-specific nature of aging. Across organ systems and molecular/cellular pathways, the biology of aging has been shown to have various, complex sex-specific patterns [12, 13]. The female reproductive system is the first organ system to age in humans, with ovarian functional decline beginning in the thirties, and the average onset of menopause in the early fifties [14]. Menopause has been shown to accelerate biological aging [15] and increase risk of multiple diseases including cardiovascular disease [16] and osteoporosis [17]. Small scale studies have also documented associations between DNAm age and implantation failure and diminished ovarian reserve [18].

Biological age estimates from existing epigenetic clocks have generally poor correlation with chronological age when applied to DNAm data collected from hormone-sensitive organs such as the endometrium, uterus, and ovaries [18, 19]. More robust aging clocks for the female reproductive system could give key insights into how accelerated reproductive aging impacts fertility, menopause, and risk of reproductive disorders such as PMOS (formerly PCOS) and endometriosis. More accurate female reproductive age estimates might also serve as important personalized biomarkers of risk. Thus, it is imperative to develop epigenetic aging clocks specific to female biology.

The exciting possibility exists to measure female reproductive age acceleration by applying clock models to DNAm data derived from menstrual fluid (MF). Recently, menstrual fluid (MF) has emerged as a potential diagnostic tool, as it is cost-effective and easy to collect, noninvasive. MF yields a unique mixture of live cells from the endometrium, cervix, vagina, blood and immune system [20] and has been shown to contain biomarkers for endometriosis, HPV/cervical cancer, and diabetes [21-23]. DNAm patterns in the endometrium in particular are complicated by the organ’s complex immune-driven regeneration process and high cell turnover rate among others. Useful clock models have the ability to separate methylation signal (the differences most strongly associated with chronological age progression), from methylation noise (caused by technical measurement artifacts) [24]. An improved epigenetic clock for use in MF would thus yield age predictions that are closer to participant chronological age than existing clocks, with lower mean absolute error (MAE) and higher linear correlation with chronological age [4]. As a first step toward gaining accurate reproductive age predictions from MF, at Xella Health we developed a novel, female-specific epigenetic aging clock based on deep learning that is optimized for performance in MF and endometrial tissue. We trained the clock on thousands of curated public female samples and validated it in MF samples from two diverse patient cohorts.

## RESULTS

### Menstrual fluid cell type composition is influenced by collection methodology

To better understand the variation in cell type composition of MF and its potential impact on the prediction of reproductive age, we first conducted cell-type deconvolution analyses of 66 MF samples collected from women between the ages of 22y and 51y (22 menstrual cup collected; 44 tampon collected; Supp. Table 1). MF is a unique sample type which contains both blood (primarily immune) and reproductive tissue resulting from the shedding of the endometrium; to estimate the broad cell type proportions of epithelial, stromal (fibroblast-like), and immune cell types in the MF samples, we applied the cell type deconvolution algorithms EpiDISH and HEpiDISH [25-27]. MF samples are primarily composed of blood immune cells with lesser proportions of epithelial and stromal cells (Figure 1B). The proportion of these cell types significantly differed between tampon and menstrual cup-collected samples (Wilcoxon rank-sum, all p<0.05), and tampon samples had higher sample-to-sample variance in cell type proportion across all 3 cell types (Figure 1B). Further deconvolution of the immune cell populations into 7 major subtypes showed a significant difference in the proportion of all immune cell sub-types between tampon and menstrual cup samples with the exception of CD8+ T-cells (Figure 1C). Principal components analysis of MF methylation data against peripheral blood, endometrial biopsies, and cervical tissue training data also showed that the methylation profile of MF samples is likely a mixture of signatures from blood and endometrial tissue (Figure 1A).

**Figure 1.**
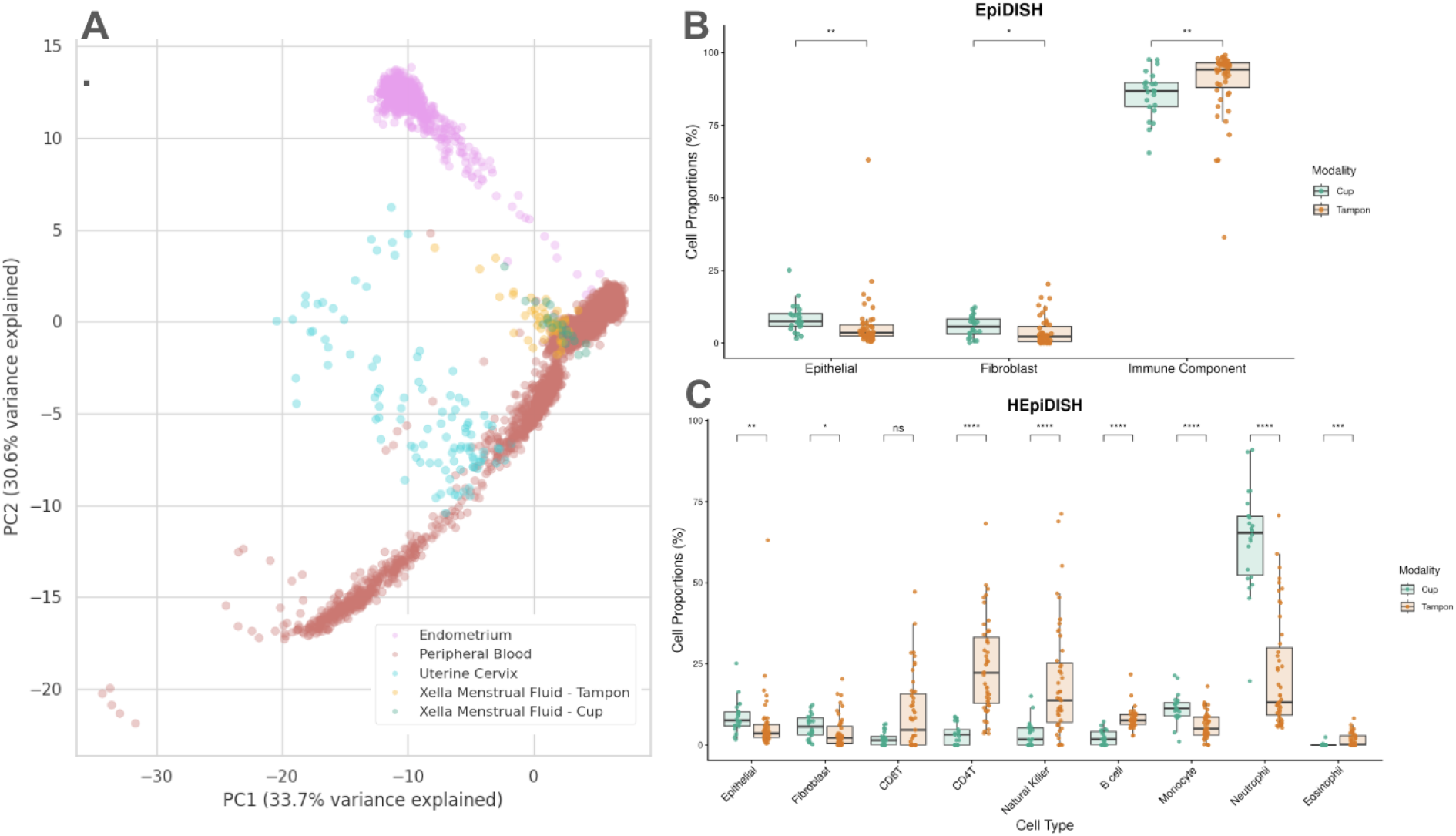
A) Principal component analysis (PCA) plot visualizing Xella MF data and publicly available methylation data from peripheral blood, endometrium, and uterine cervix samples. B) Estimated proportion of epithelial cells, stromal (fibroblast-like), and immune cells in each MF sample using EpiDISH. C). Estimated proportion of immune cell subtypes in each MF sample (HEpiDISH). *p<0.05,**p<0.01, ***p<0.001, n.s. p>0.05.

Gynecological disease leading to inflammation and baseline hormone levels have the potential to impact the cell type composition and thus the methylation profile of menstrual fluid samples. However, we found no significant differences in estimated cell type proportions between MF samples from patients with endometriosis and healthy controls (Supp. Figure 1). The use of hormonal contraceptives is also known to change the composition and thickness of the endometrium [28] and we sought to determine whether hormonal contraceptives influence the estimated cell type proportions of menstrual fluid. In a small subset of participants using hormonal contraceptive methods (n=8), we did not find any significant differences in the proportion of cell subtypes versus participants not using birth control (n=20) (Supp. Figure 2), however, more data is necessary to examine this question with greater power.

**Figure 2.**
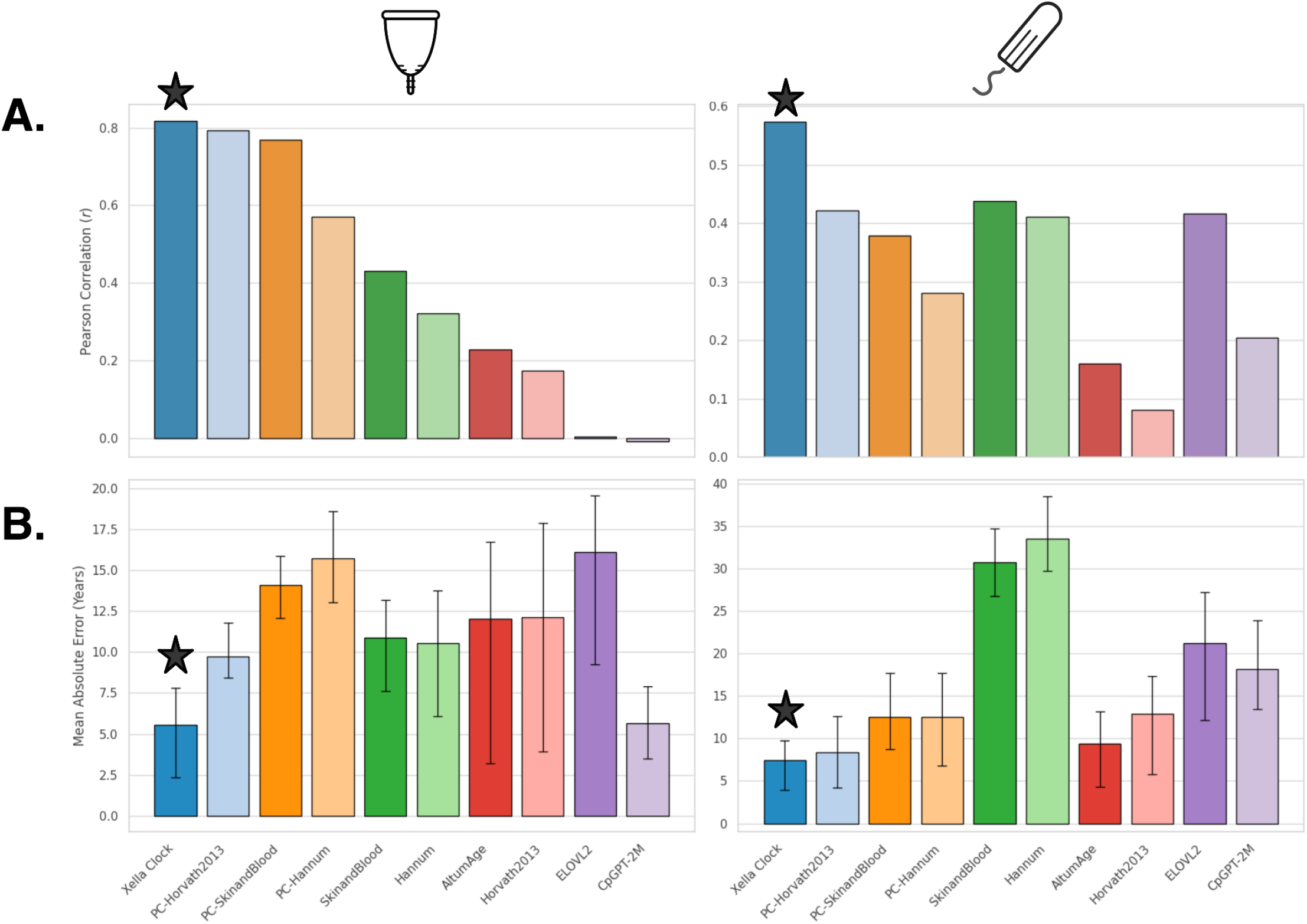
Performance of epigenetic aging clocks on MF samples, Xella Clock shown in dark blue. A) Pearson correlation between predicted age and participant chronological age in menstrual cup MF data (left) and tampon MF data (right). B) Mean Absolute Error (MAE) between predicted age and chronological age in menstrual cup MF data (left) and tampon MF data (right). Stars indicate best-performing clocks for each measure of performance. Error bars represent the inter-quartile range of absolute error values, where the upper limit is the 75th percentile and the lower limit is the 25th percentile.

### Development and Performance of The Xella MF Clock

We assessed the performance of several well-known first generation epigenetic aging clocks including Hannum [29], Horvath (2013) [4], the Skin and Blood clock [30], their corresponding Principal Component (PC)-based clocks [24], AltumAge [31], ELOVL2 [32], and CpGPT-2M age for predicting chronological age from MF samples (Figure 2). ELOVL2 is the simplest clock (3-markers) and is often used for age prediction in degraded or contaminated biological sample types. CpGPT-2M age is the largest model based on deep learning and trained on almost all available public methylation data across tissue types. We observed that existing clocks struggled to accurately predict age from MF data particularly for tampon-collected samples. Among the existing clocks, the PC-Horvath clock yielded the best age correlation for menstrual cup collected samples (Pearson r=0.79, Figure 2A, left), yet had a high MAE of 9.7 years (Figure 2B, right). In tampon-collected samples, the Skin and Blood clock yielded the highest correlation value of 0.44 (Figure 2A, right), but also had an extremely high MAE of 30.7 years (Figure 2B, right). The PC-Horvath clock had a much lower MAE of 8.4 years in tampon-collected samples (Figure 2B, right), with a correlation value of 0.42, (Figure 2A, right).

Given the limited performance of existing epigenetic age clocks for predicting chronological age from MF data, particularly for tampon-derived samples, we developed a novel, female-specific aging clock optimized for performance in MF and endometrial tissue (see Methods for further details). We trained and tested several major clock model architectures including classical elastic-net regression models, boosted trees, and custom principal components based models. We trained each model on a variety of public data sets filtered by sex, age, and tissue type. All MF samples were reserved as a held-out test set. We found an optimized DNN-based clock had the best performance in both tampon and cup held-out test samples. The best performing model, the Xella Clock, was trained on 7,052 publically available female samples from peripheral blood, endometrium, and cervix (see Methods).

The Xella Clock outperformed existing epigenetic aging clocks for age prediction in both tampon and MC collected samples. Age predictions from our novel clock achieved a simultaneous increase in chronological age correlation and a decrease in prediction error (MAE) (Figure 2, marked by star). In menstrual cup collected samples, age correlation increased to r=0.82 and MAE was reduced to 5.5y (Figure 3B). In tampon-collected samples, age correlation increased to 0.58 and MAE decreased to 7.4y (Figure 3A).

**Figure 3.**
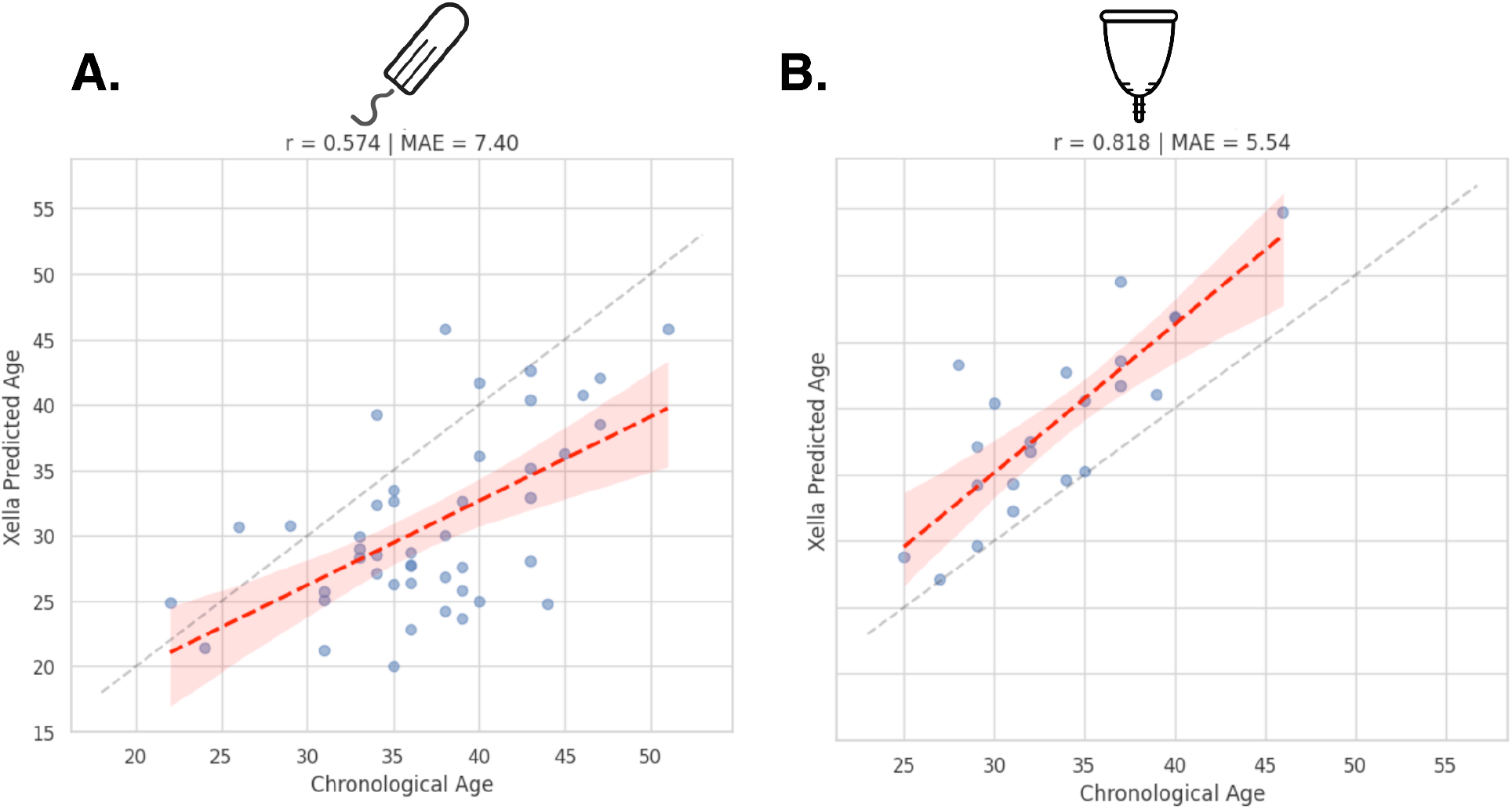
Scatterplots showing the correlation between chronological age and age prediction from the Xella Clock using **A)** tampon collected MF samples and **B)** menstrual cup samples.

Most existing epigenetic clocks, as well as the Xella Clock, had greater error of age prediction in tampon-collected vs. menstrual cup samples. Although tampon and cup collected samples differed significantly in immune cell subtype proportion (Fig. 1), we found this factor alone could not explain the different performance of the Xella Clock between the two sample types. The prediction performance of the Xella Clock was not improved by the inclusion of epithelial and stromal cell type proportion information from training samples in Xella Clock model training (data not shown). Immune cell type proportions were also not a significant predictor of absolute age error of the clock for any cell type (Supp. Table 2). Tampon and cup collected samples also did not differ significantly in sequencing quality or median filtered site-specific sequencing coverage (Supp. Figure 3).

### Validation of the Xella Clock on external MF and endometrial data

We conducted an additional validation of the Xella Clock on an independent menstrual effluent dataset (GSE275888) containing 12 patients, each who provided menstrual cup, pad, and vaginal swab samples (n=36 samples) [34]. The Xella Clock predictions showed high correlation with chronological age and low MAE in all 3 sample types, with the best overall performance on menstrual cup samples (Figure 4).

**Figure 4.**
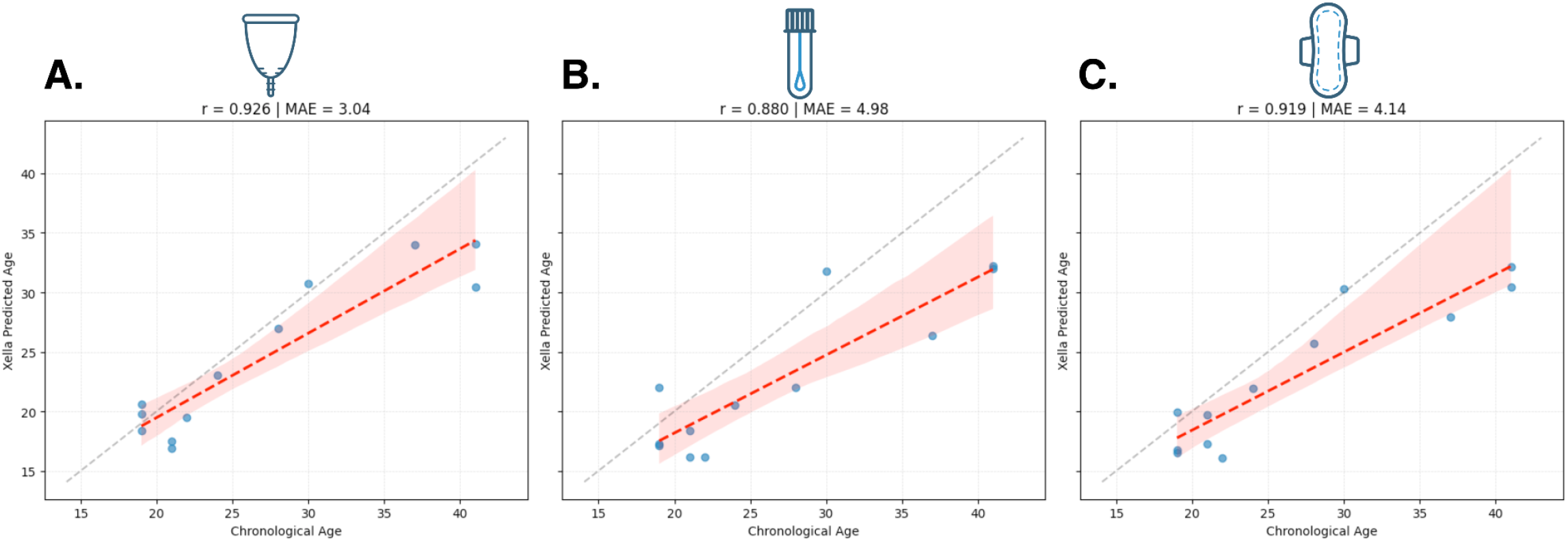
Correlation between Xella Clock predicted ages (y axis) and chronological ages (x axis) of menstrual fluid samples from GSE275888. A) menstrual cup samples B) vaginal swab samples, and C) pad samples.

To further assess the reliability of our clock, we tested it on an external endometrial methylome dataset (GSE90060) that was not included in model training, validation, or previous testing. This dataset included endometrial tissue from females in early secretory and mid-secretory phases of the menstrual cycle (17 patients, 34 samples) [35]. Despite biological differences in the composition of the endometrium in these phases, our clock achieved high age correlation and low MAE in samples from both phases (Figure 5).

**Figure 5.**
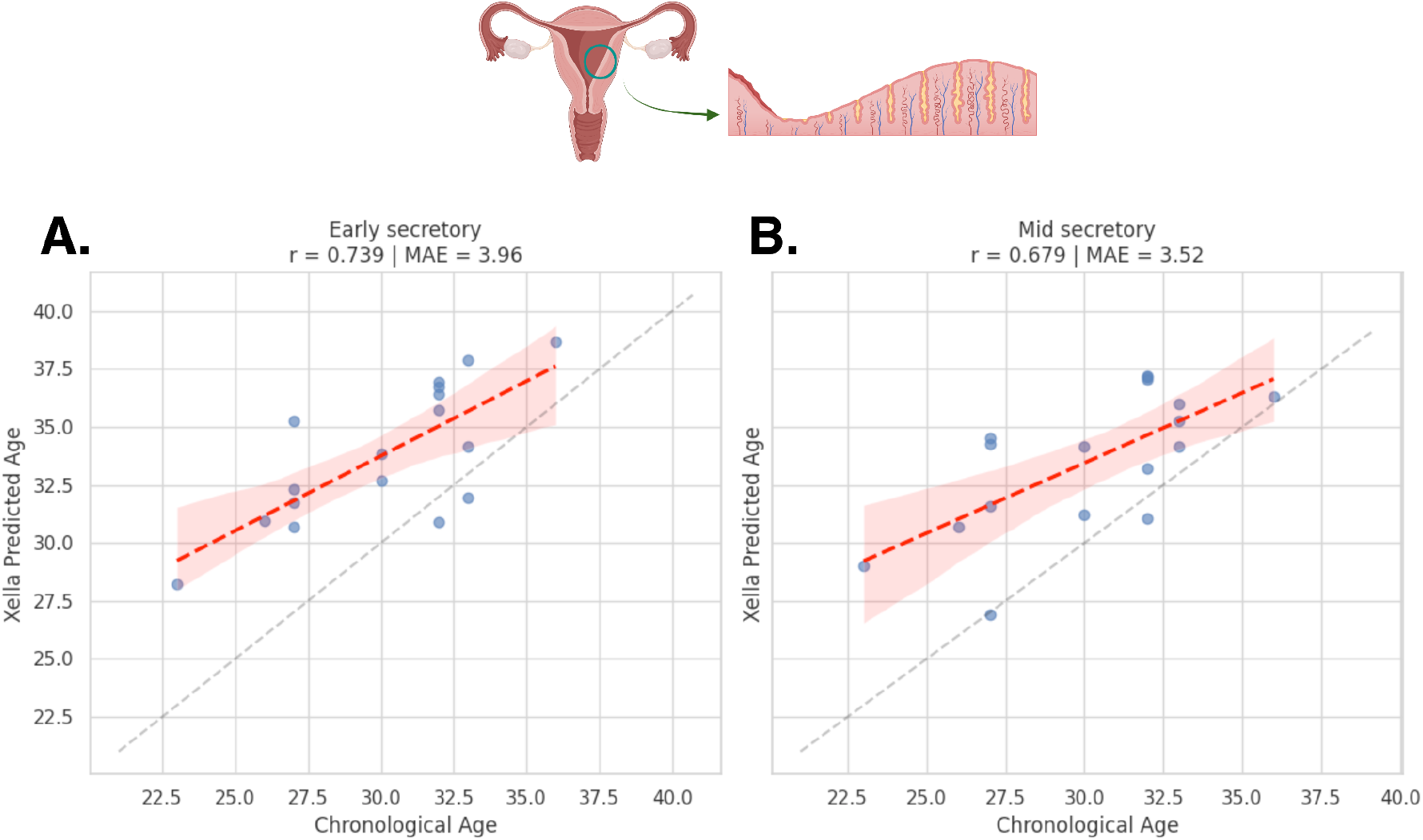
Scatterplots showing correlation between Xella Clock predicted age (Y axis) and chronological age (x-axis) in endometrial tissue samples from an external endometrial methylome dataset (GSE90060). Results are separated by the A) early secretory and B) mid secretory menstrual cycle phases.

## DISCUSSION

The Xella Clock is a novel deep learning-based epigenetic aging clock specific to female biology that is optimized for performance in menstrual fluid and endometrial tissue samples. The clock outperforms existing clocks at chronological age prediction from menstrual fluid and endometrial tissue by more effectively leveraging biological methylation signal that is correlated with chronological age progression in MF. Performance of the Xella Clock model was validated in an external MF data set and also in an external endometrial tissue data set. This model can be used to more accurately test the association between MF age acceleration and health factors, such as infertility and risk of gynecological disease, in ongoing large scale studies.

While the Xella Clock has improved age prediction performance in MF samples, we observed marked differences between the efficacy of the clock in samples collected using different modalities (menstrual cup vs. tampon). There was lower accuracy in tampon collected samples, and these samples also had high variability in endometrial and immune cell type proportions. Tampon derived samples are also likely subject to greater sample degradation during collection. Xella Clock prediction results for tampon-collected MF samples from a separate study (Fig 4) had a similar level of accuracy to that observed for our own collections.

Further studies are needed to understand how clinical factors may impact MF age predictions. One important consideration moving forward is how MF and endometrial epigenetic age predictions may vary in patients using hormonal contraceptives or hormone replacement therapies. Hormone exposure is known to impact cycle regularity as well as thickness of the endometrial lining, and this in turn may impact age acceleration measurements made by epigenetic models. Another potential area for optimization of the clock includes integration of patient cycle-phase information for improved age acceleration measurements in samples derived from non-menstrual phase endometrial tissue biopsies. We found that self-reported endometriosis diagnosis status was not significantly associated with the Xella Clock’s age prediction error. This is consistent with other studies in which eutopic endometrial tissue from patients with endometriosis does not show an accelerated biological age [36] (Supp. Figure 4).

### Future Directions

Using the Xella Clock as a starting point, we see great potential for understanding the associations between endometrial reproductive age acceleration in MF, infertility, and common gynecological diseases such as PMOS, leiomyomas (fibroids), and endometriosis. Measurements of MF age acceleration in turn can be used to provide a personalized method of disease monitoring and fertility planning for individual patients. MF age acceleration measurements can be easily tracked longitudinally over time in order to detect biological changes throughout a woman’s reproductive years. With the addition of parallel multiomic measurements in MF, there is also the potential to detect complex multiomic biomarkers of inflammation and gynecological disease risk prior to the onset of symptoms. Through the development of this and additional multiomic MF assays, we aim to greatly increase the number of disease screening and overall health optimization tools available to women in the near future.

## Data Availability

Summary statistics for data produced in the present study are available upon reasonable request to the authors.

## Acknowledgements

The authors gratefully acknowledge Dr. Sally Mortlock and Dr. Irma Vlasac for their support of this work.

## METHODS

### Xella menstrual fluid data collection

Menstrual fluid (MF) samples were obtained from a Xella Health IRB approved health study volunteers using FDA-approved tampons or menstrual cup, preserved with DNA Shield buffer (Zymo Research, Inc.) and sequenced using HiFi long-reads (Revio platform, PacBio Inc.). MF samples were sequenced from 9.4X to 44.6X genome-wide filtered mean read coverage (Supp. Figure 3; menstrual cup average, 23.9X; tampon average, 26.3X). Methylation calls were measured as model-derived scores using pb-cpg-tools [37]. Methylation calls at sites within 5bp of an SNV were filtered out. Samples with greater than 75% missing methylation calls across the set of sites included on the EPICv2 Illumina methylation array were also excluded. Remaining missing CpG calls were imputed as the cohort mean value for that site. Quality filtering retained data from 66 participants, 44 of whom provided samples collected via tampon and 22 of whom provided samples collected via menstrual cup (Figure 6). Clinical characteristics of the participant cohort are provided in Supplementary Table 1.

**Figure 6.**
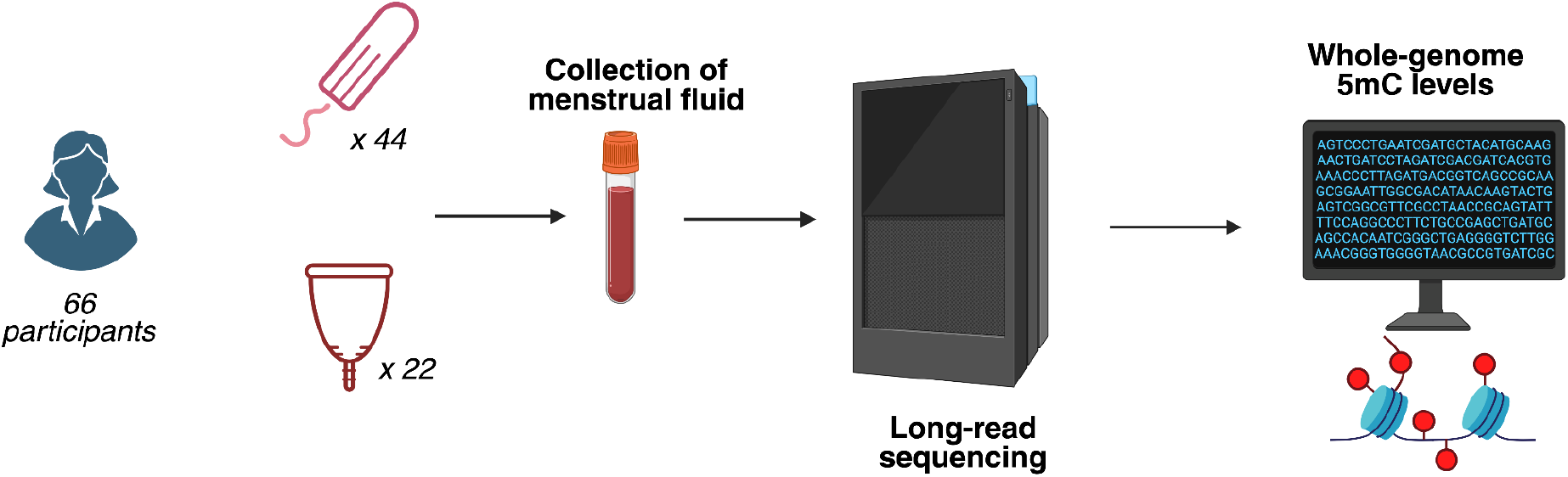
Schematic overview of Xella menstrual fluid collection and sequencing.

### Model Development and Training Data

To develop a novel clock, we curated a dataset of publicly available methylation data composed of only female samples spanning three primary tissue types: peripheral blood, endometrial tissue, and uterine cervix tissue, from the NCBI GEO database (https://www.ncbi.nlm.nih.gov/geo/, [37]), the CpGPT S3 repository [33], and the AltumAge repository [31]. The final body of training data consisted of 7,052 samples, which we split into training, validation, and test sets (80:10:10).

We developed and tested models based on different machine learning methodologies including elastic net regression, random forest regressors, XGBoost regressors, custom principal components-based models, as well as deep neural networks (DNNs). Due to its relatively small sample size, Xella MF data was not used for training the models but instead used as an additional held-out test set. For a subset of models we evaluated the use of MF data in a cross-validation framework. Age predictions from published clock models were generated with the pyaging library [39]. Novel machine learning models were developed using the Scikit-learn library in Python [40], and DNNs were designed with pytorch [41] and optimized with the Optuna library [42]. Cell type proportion analysis was performed using EpiDISH and HEpiDISH software [25-28]. Multiple linear regression analysis on Xella Clock age prediction error was performed using the proportion of each cell type and the collection methodology as predictors with the statsmodels package.

Ultimately, an optimized DNN model trained solely on public data outperformed all other models for age prediction in MF samples, and we chose this architecture for the Xella Clock. Loss curves and performance on training, validation, and test data for the Xella Clock are provided in Figure 7.

**Figure 7.**
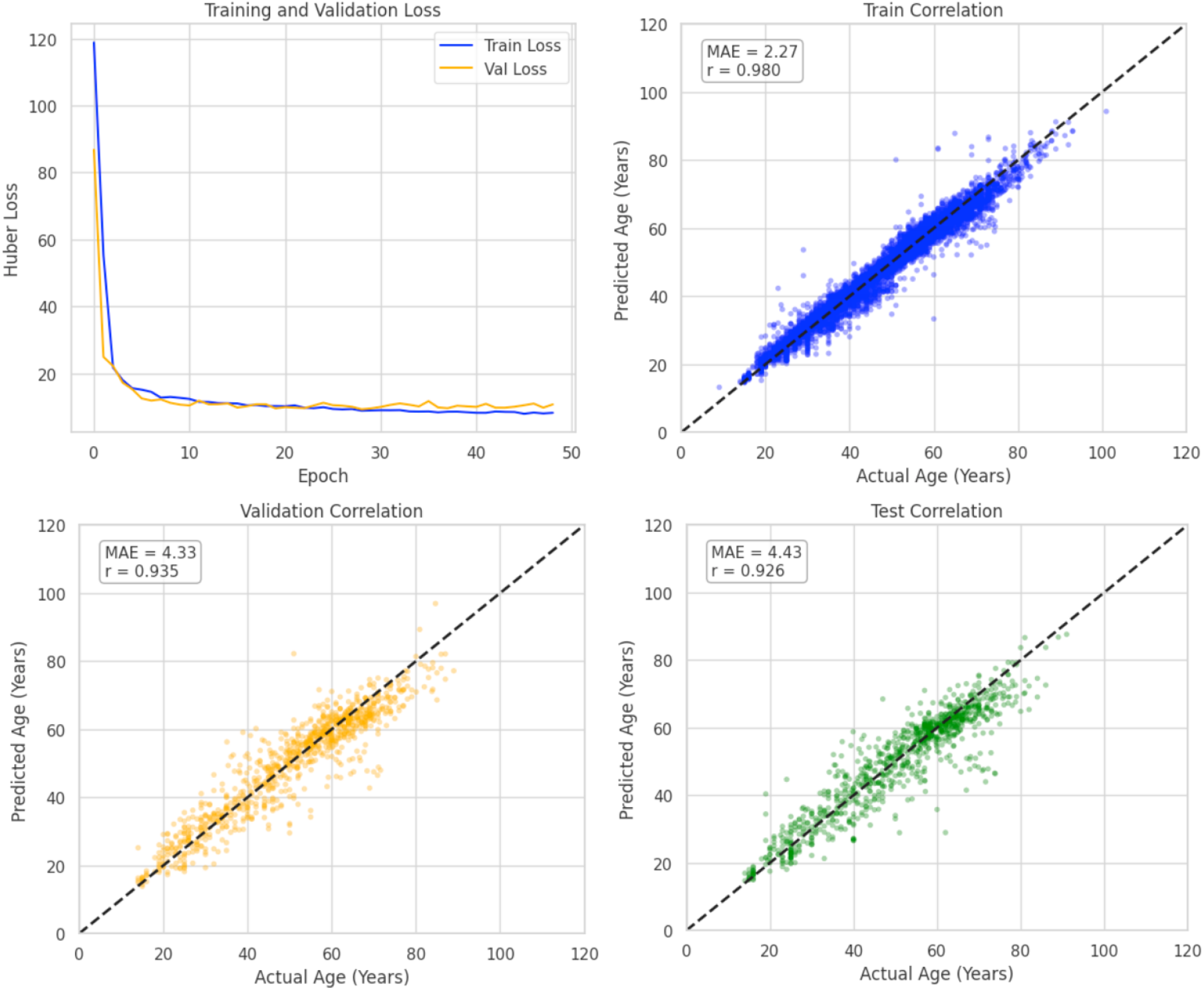
Loss curves (using PyTorch Huber Loss) and scatterplots of Pearson correlation between model-predicted age and actual age in the training data, validation data, and test data. Training with early stopping was used with a patience of twenty iterations for validation loss improvement. Xella menstrual fluid data was not included in the train, validation, or test set.

## SUPPLEMENTARY INFORMATION

**Supplementary Table 1.** Clinical characteristics of Xella Health study volunteers.

| Characteristic | n | % |
| --- | --- | --- |
| **Collection Method** |  |  |
| Tampon | 44 | 66.7% |
| Menstrual Cup | 22 | 33.3% |
| **Age** |  |  |
| N | 66 |  |
| Mean (SD) | 35.8 (5.95) |  |
| Median | 35.5 |  |
| Range | 22.0, 51.0 |  |
| **Endometriosis Status** |  |  |
| Endometriosis | 21 | 31.8% |
| Control | 45 | 68.2% |
| **Race/Ethnicity** |  |  |
| White (Non-Hispanic) | 24 | 36.4% |
| Non-White | 20 | 30.3% |
| Unknown | 22 | 33.3% |
| **Birth Control Method** |  |  |
| Hormonal | 8 | 12.1% |
| Non-hormonal | 15 | 22.7% |
| None | 20 | 30.3% |
| Unknown | 23 | 34.8% |

**Supplementary Table 2.**
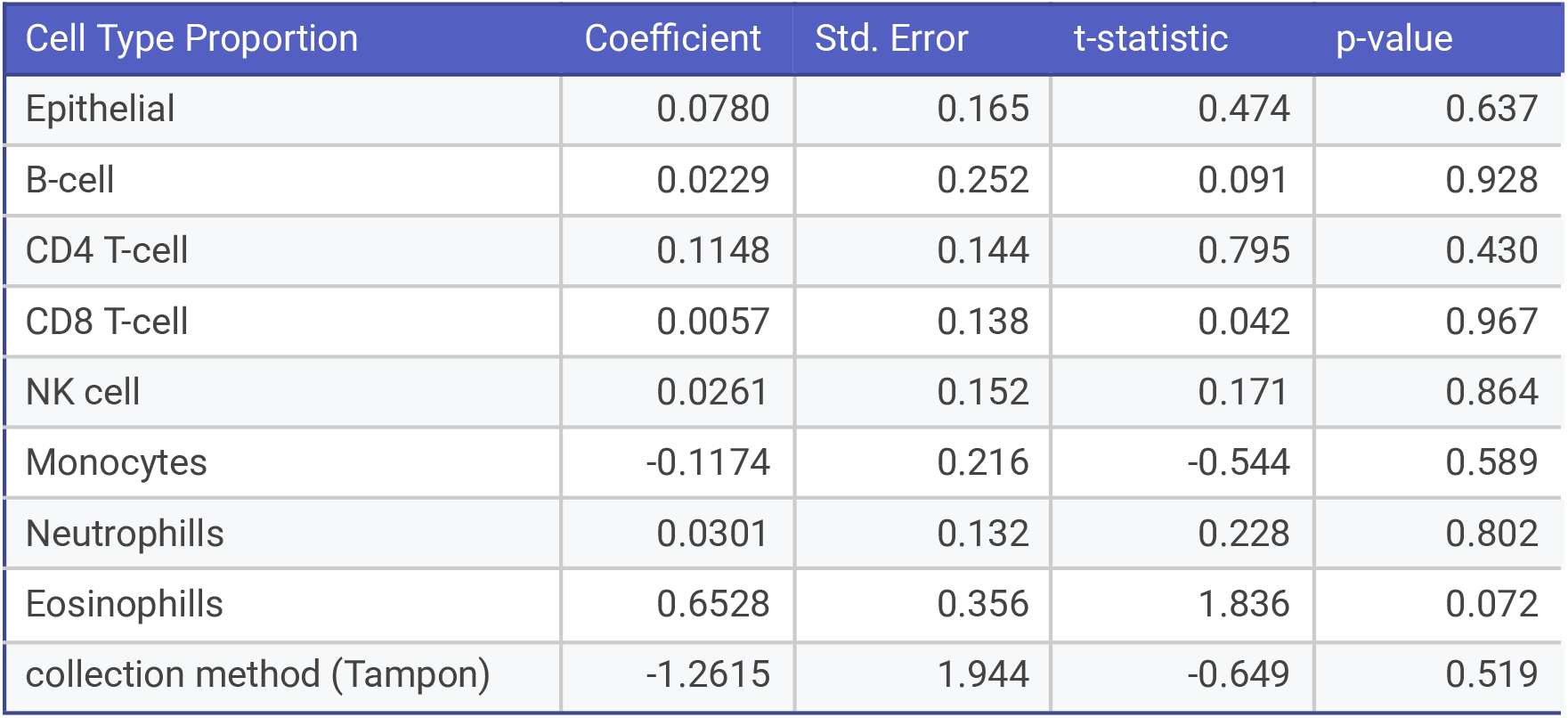
Multiple linear regression analysis of Xella Clock absolute error against the sample cell type proportions and collection method (Rsq adj.=0.071). Fibroblast-like cell type and menstrual-cup collection method were set as the reference classes in the analysis.

| Cell Type Proportion | Coefficient | Std. Error | t-statistic | p-value |
| --- | --- | --- | --- | --- |
| Epithelial | 0.0780 | 0.165 | 0.474 | 0.637 |
| B-cell | 0.0229 | 0.252 | 0.091 | 0.928 |
| CD4 T-cell | 0.1148 | 0.144 | 0.795 | 0.430 |
| CD8 T-cell | 0.0057 | 0.138 | 0.042 | 0.967 |
| NK cell | 0.0261 | 0.152 | 0.171 | 0.864 |
| Monocytes | -0.1174 | 0.216 | -0.544 | 0.589 |
| Neutrophils | 0.0301 | 0.132 | 0.228 | 0.802 |
| Eosinophils | 0.6528 | 0.356 | 1.836 | 0.072 |
| collection method (Tampon) | -1.2615 | 1.944 | -0.649 | 0.519 |

**Supplementary Figure 1.**
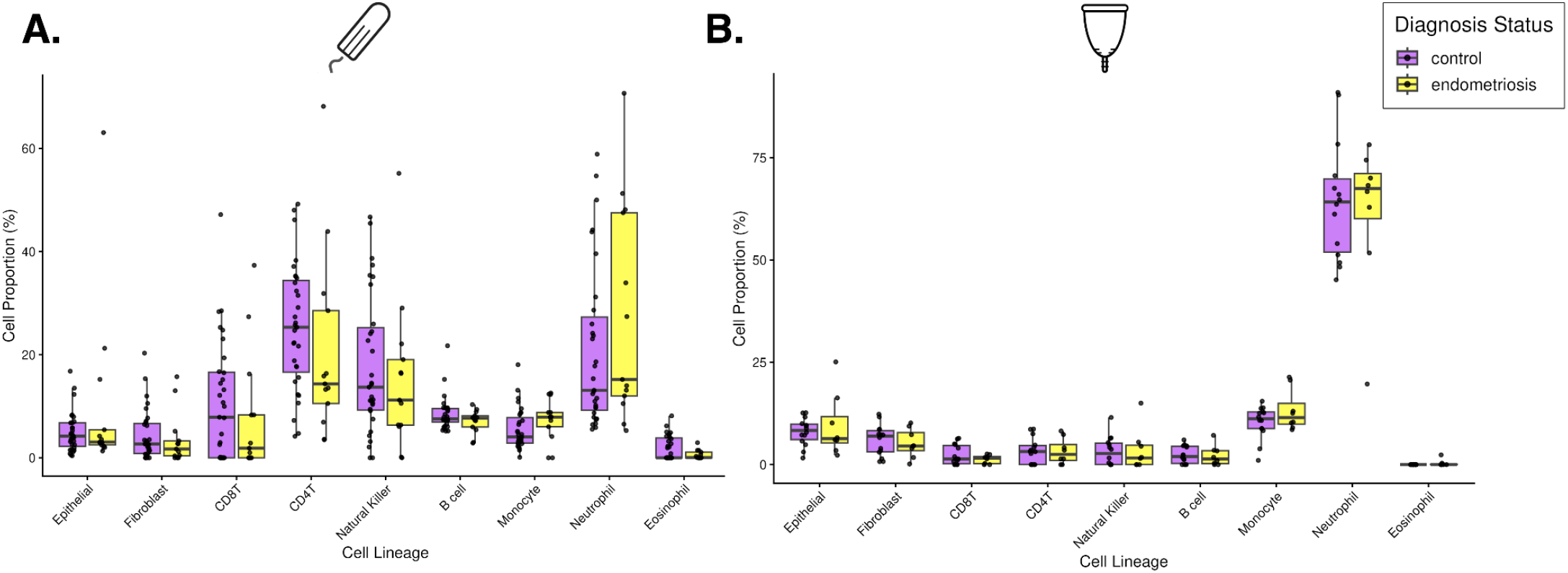
Comparison of estimated cell type proportions in healthy participants (purple) vs those with endometriosis (yellow) in A) tampon-collected MF samples, B) menstrual cup-collected samples. The difference in cell-type proportion between cases and controls was not statistically significant (p>0.05) for any cell type.

**Supplementary Figure 2.**
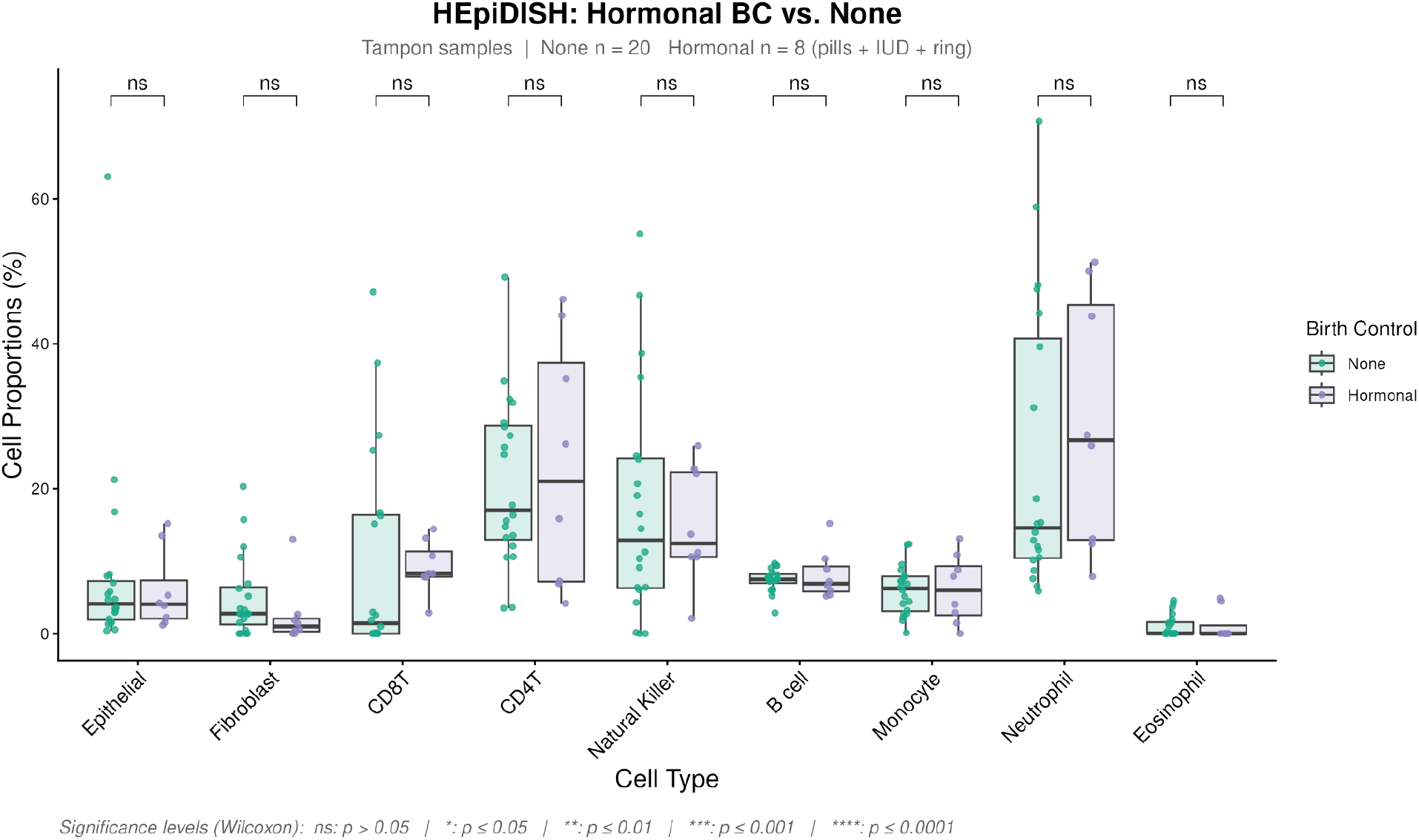
Comparison of estimated cell type proportions in MF samples from participants using hormonal birth control (light purple) and participants not using birth control (blue-green). Cell type proportions were estimated using HEpiDISH. All samples included in this figure are tampon-collected.

**Supplementary Figure 3:**
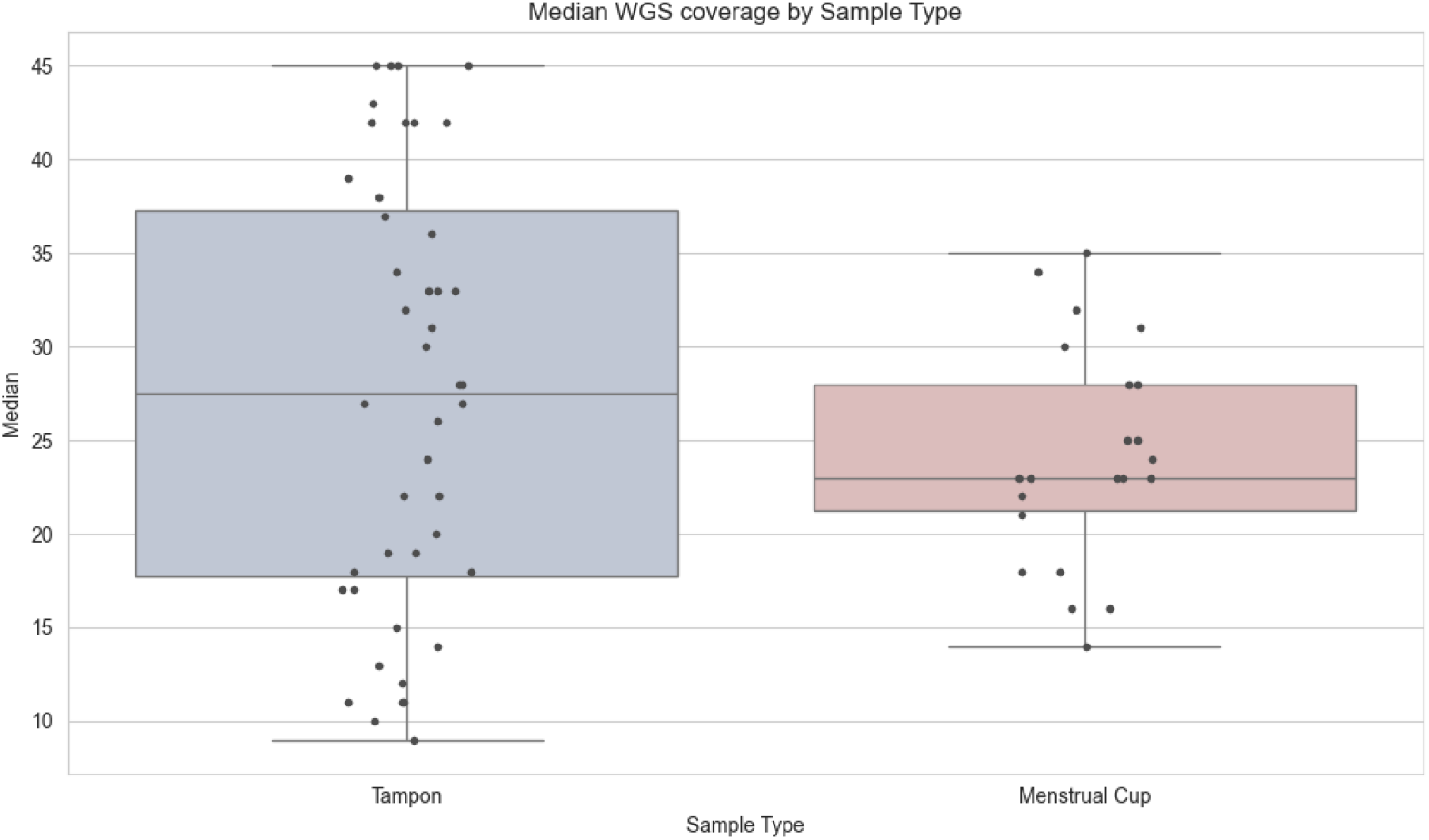
Median WGS coverage by sample type. Coverage did not differ significantly between sample types (Welch’s t-test, p = 0.23).

**Supplementary Figure 4.**
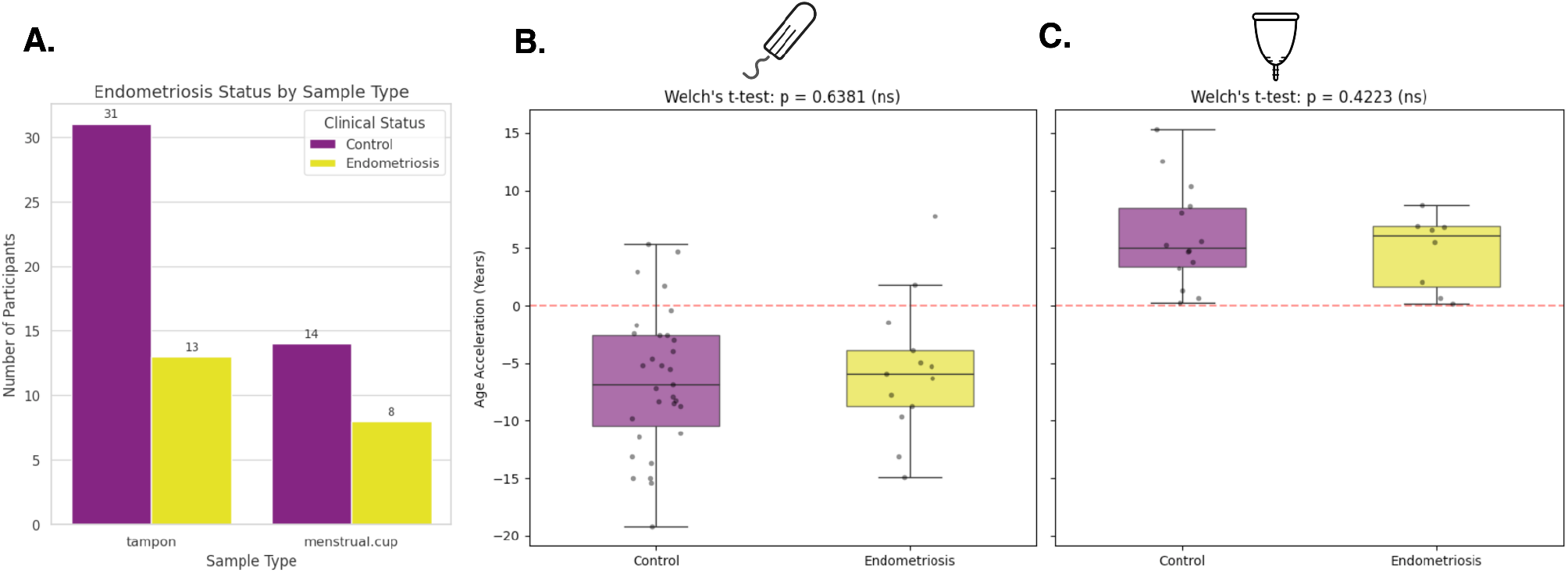
A) Number of participants with endometriosis (yellow) and without endometriosis (purple) in both tampon and menstrual cup modalities. B) Age acceleration/deceleration in years, quantified by the residuals between Xella Clock prediction and chronological age. No significant difference in age acceleration between control and endometriosis (Welch’s t-test, p > 0.05). C) Age acceleration/deceleration in years in menstrual cup samples (Welch’s t-test, p > 0.05).

## REFERENCES

1. López-Otín, C., Blasco, M. A., Partridge, L., Serrano, M. & Kroemer, G. Hallmarks of aging: An expanding universe. Cell 186, 243–278 (2023).

2. Seale, K., Horvath, S., Teschendorff, A. et al. Making sense of the aging methylome. Nat Rev Genet 23, 585–605 (2022).

3. Yang, X., Shao, X., Gao, L., Zhang, S. Comparative DNA methylation analysis to decipher common and cell type-specific patterns among multiple cell types, Briefings in Functional Genomics, Volume 15, Issue 6, Pages 399–407 (2016)

4. Horvath, S. DNA methylation age of human tissues and cell types. Genome Biol 14, 3156 (2013)

5. Rutledge, J., Oh, H. & Wyss-Coray, T. Measuring biological age using omics data. Nat Rev Genet 23, 715–727 (2022).

6. Levine M, Lu A, Quach A, Chen B, Assimes T, Bandinelli S, Hou L, Baccarelli A, Stewart J, Li Y, Whitsel E, Wilson J, Reiner A, Aviv A, Lohman K, Liu Y, Ferrucci L, Horvath S. An epigenetic biomarker of aging for lifespan and healthspan. Aging (Albany NY). 10:573–591 (2018)

7. Lu A, Quach A, Wilson J, Reiner A, Aviv A, Raj K, Hou L, Baccarelli A, Li Y, Stewart J, Whitsel E, Assimes T, Ferrucci L, Horvath S. DNA methylation GrimAge strongly predicts lifespan and healthspan. Aging (Albany NY). 11:303–327. (2019)

8. Zalesky, A., Wen, J. & Tian, Y.E. From whole-body to organ-specific biological age clocks. Nat Aging 6, 961–969 (2026).

9. Oblak, L., van der Zaag, J., Higgins-Chen, A. T., Levine, M. E. & Boks, M. P. A systematic review of biological, social and environmental factors associated with epigenetic clock acceleration. Ageing Research Reviews 69, 101348 (2021).

10. Li, G., Cheng, L., Wong, I.N. et al. Predicting healthspan and disease risks through biological age. Trends in Molecular Medicine 32, 354–369 (2026).

11. Guo, J., Huang, X., Dou, L. et al. Aging and aging-related diseases: from molecular mechanisms to interventions and treatments. Sig Transduct Target Ther 7, 391 (2022).

12. Hägg, S. and Jylhävä, J. Sex differences in biological aging with a focus on human studies eLife 10:e63425, (2021).

13. Reicher, L., Bar, N., Godneva, A. et al. Phenome-wide associations of human aging uncover sex-specific dynamics. Nat Aging 4, 1643–1655 (2024).

14. Gold EB. The timing of the age at which natural menopause occurs. Obstet Gynecol Clin North Am. Sep;38(3):425–40. (2011).

15. Levine, M. E. et al. Menopause accelerates biological aging. Proceedings of the National Academy of Sciences 113, 9327–9332 (2016).

16. Uddenberg, E. R. et al. Menopause transition and cardiovascular disease risk. Maturitas 185, 107974 (2024).

17. de Villiers, T. J. Bone health and menopause: Osteoporosis prevention and treatment. Best Practice & Research Clinical Endocrinology & Metabolism, 38, 101782 (2024).

18. Piani, L.L, Vigano, P. and Somigliana E. Epigenetic clocks and female fertility timeline: A new approach to an old issue? Front. Cell Dev. Biol. 11:1121231 (2023).

19. Deryabin, P.I. and Borodkina, A.V. Epigenetic clocks provide clues to the mystery of uterine ageing, Human Reproduction Update, Volume 29, Issue 3, Pages 259–271, (2023)

20. Schwalie, P.C. et al., “Single-Cell Characterization of Menstrual Fluid at Homeostasis and in Endometriosis,” preprint, medRxiv, 2024.05.06.24306766, May 6, (2024).

21. Zaheer, A, Komel, A, Abu, B, et al. Potential for and challenges of menstrual blood as a non-invasive diagnostic specimen: current status and future directions. Annals of Medicine & Surgery 86(8):p 4591–4600, August 2024

22. Warren, L.A., Shih, A., Renteira, S.M. et al. Analysis of menstrual effluent: diagnostic potential for endometriosis. Mol Med 24, 1 (2018).

23. Chakravarti P, Maheshwari A, Tahlan S, et al. Diagnostic accuracy of menstrual blood for human papillomavirus detection in cervical cancer screening: a systematic review. Ecancermedicalscience. Jul 14;16:1427.(2022).

24. Higgins-Chen, A, et al., A Computational Solution for Bolstering Reliability of Epigenetic Clocks: Implications for Clinical Trials and Longitudinal Tracking. Nature Aging 2, no. 7 : 644–61, (2022).

25. Teschendorff, A.E., Breeze, C.E., Zheng, S.C., and Beck, S. A comparison of reference-based algorithms for correcting cell-type heterogeneity in Epigenome-Wide Association Studies. BMC Bioinformatics 18 (1):105. (2017).

26. Zheng, S.C., Breeze, C.E., Beck, S., and Teschendorff, A.E. Identification of differentially methylated cell-types in Epigenome-Wide Association Studies. Nature Methods 15 (12):1059–66. (2018).

27. Zheng, S.C., Webster, A.P., Dong, D., et al. A novel cell-type deconvolution algorithm reveals substantial contamination by immune cells in saliva, buccal and cervix. Epigenomics 10 (7):925–40. (2018)

28. Talukdar, N., Bentov, Y, Chang, P.T., et al. Effect of Long-Term Combined Oral Contraceptive Pill Use on Endometrial Thickness. Obstetrics & Gynecology 120(Part 1):p 348–354. (2012).

29. Hannum G, Guinney J, Zhao L, Zhang L, et al. Genome-wide methylation profiles reveal quantitative views of human aging rates. Mol Cell, 49:359–367. (2013)

30. Horvath S, Oshima J, Martin G, Lu A, et al. Epigenetic clock for skin and blood cells applied to Hutchinson Gilford Progeria Syndrome and ex vivo studies. Aging (Albany NY). 10:1758–1775. (2018).

31. de Lima Camillo, L.P., Lapierre, L.R. & Singh, R. A pan-tissue DNA-methylation epigenetic clock based on deep learning. npj Aging 8, 4 (2022).

32. Garagnani, P et al., “Methylation of ELOVL2 Gene as a New Epigenetic Marker of Age,” Aging Cell 11, no. 6: 1132–34. (2012).

33. de Lima Camillo, L. P. et al. CpGPT: a Foundation Model for DNA Methylation. bioRxiv 2024.10.24.619766 (2025).

34. Vlasac IM, Stolrow HG, Thayer ZM, Christensen BC, Rivera L et al. DNA-based cell typing in menstrual effluent identifies cell type variation by sample collection method: toward noninvasive biomarker development for women’s health. Epigenetics. 20(1):2453275. (2025).

35. Kukushkina, V., Modhukur, V., Suhorutšenko, M. et al. DNA methylation changes in endometrium and correlation with gene expression during the transition from pre-receptive to receptive phase. Sci Rep 7, 3916 (2017).

36. Leap, K., Yotova, I., Horvath, S. et al. Epigenetic age provides insight into tissue origin in endometriosis. Sci Rep 12, 21281 (2022).

37. Mortlock, S. et al., Global Endometrial DNA Methylation Analysis Reveals Insights into mQTL Regulation and Associated Endometriosis Disease Risk and Endometrial Function. Communications Biology 6, no. 1:p780, (2023).

38. PacificBiosciences/Pb-CpG-Tools, Rust, March 21, 2022; PacBio, released July 8, 2026, https://github.com/PacificBiosciences/pb-CpG-tools.

39. de Lima Camillo, L. P. et al. pyaging. Version 0.1.30. https://pypi.org/project/pyaging/ (2024).

40. Pedregosa, F. et al. Scikit-learn: Machine Learning in Python. J. Mach. Learn. Res. 12, 2825–2830 (2011).

41. Chintala, S. et al. PyTorch. Version 1.0.2. https://pypi.org/project/pytorch/ (2017).

42. Akiba, T. et al. Optuna. Version 4.9.0. https://pypi.org/project/optuna/ (2018).

